# OCR-Mediated Modality Dominance in Vision-Language Models: Implications for Radiology AI Trustworthiness

**DOI:** 10.64898/2026.02.22.26346828

**Authors:** Izzet T. Akbasli, Baris Ozturk, Oguzhan Serin, Volkan Dogan, Goksu Bozdereli Berikol, Donnella S. Comeau, Leo A. Celi, Orhan Ozguner

## Abstract

1.

**Background:** Vision-language models (VLMs) are increasingly proposed for radiologic decision support, yet the security implications of deploying general-domain, OCR-capable models in diagnostic workflows remain poorly characterized. When image-embedded text is not treated as untrusted input, the visual channel becomes vulnerable to adversarial manipulation through OCR-readable overlays.

**Methods:** Nine commercial VLMs, none intended or validated for clinical diagnosis, were evaluated on 600 brain MRI studies (300 tumor-positive, 300 tumor-negative) for binary tumor detection across four conditions: clean input, visible radiology-report injection, human-imperceptible stealth OCR injection, and a multi-stage immune-prompt defense combining both attack types with enforced visual-priority reasoning. Approximately 27,000 inference calls were analyzed. Primary outcomes included accuracy, attack success rate (ASR), false positive rate (FPR), and masking rate.

**Results:** At baseline, performance was heterogeneous (median accuracy 0.69, sensitivity 0.79, specificity 0.59). Visible injection caused universal specificity collapse (0.00 across all models; FPR 1.00), with a median ASR of 0.97; every model unconditionally privileged the injected text over its own visual analysis. Stealth injection, despite being imperceptible to human reviewers, still drove substantial degradation (median accuracy 0.43; ASR 0.57; FPR 0.84). Immune prompting achieved only partial and inconsistent mitigation: under stealth injection, median ASR decreased to 0.44, and accuracy improved to 0.56, yet residual overcalling persisted (median FPR 0.67), and three models maintained an FPR of 1.00.

**Conclusions:** Commercial VLMs exhibit a deployment-critical failure mode in radiology-like scenarios: OCR-readable text embedded in images can dominate the decision pathway and override pixel-level evidence, even under stealth conditions that evade human inspection. Prompt-level defenses provide insufficient protection. These findings underscore that any clinical integration of VLMs must be gated by system-level safeguards, including OCR-aware input handling, provenance controls, and enforced human verification, before such tools can be considered for safety-sensitive environments.

## 2. Introduction

### Background and Motivation

Large language models (LLMs) and vision-language models (VLMs) are increasingly integrated into clinical software systems, including electronic health records, clinical information extraction, and decision-support tooling ^1–7^. Unlike text-only LLMs, VLMs jointly process images and language, introducing a second input channel that carries pixel-level clinical evidence and may also contain text readable through optical character recognition (OCR)^2–4^.

In medical settings, language models may reduce documentation burden and support guideline-oriented workflows^8–10^, while multimodal extensions enable report-aware reasoning in radiology and related domains^11–13^. However, clinical deployment can amplify automation bias^14,15^. Multi-model and multi-agent safeguards have been proposed to mitigate this tendency^16–22^, but these mitigations assume that the input itself has not been compromised.

A less characterized risk arises from the multimodal attack surface itself. Many VLMs perform OCR as a first-class capability, allowing text embedded within images to influence outputs^11–13,23^. If imaging workflows permit untrusted overlays to enter the model input channel, OCR-readable content can act as an instruction vector that overrides pixel-level evidence^3,23,24^. Radiology is particularly susceptible because images routinely contain overlays that can camouflage injected content, and identifying information may persist as burned-in pixel text, creating coupled privacy and safety risks^25,26^. In this context, the central problem is deployment governance: a system-level failure mode in which nonvalidated outputs are consumed as diagnostic-grade evidence without controls for input provenance, OCR-aware sanitization, and escalation to human verification^14,15^.

### Problem Statement

In this work, we evaluate commercial general-domain VLM endpoints that are neither radiology-trained nor clinically validated, to characterize a deployment governance risk rather than claim diagnostic capability^3,11–13^. Our threat model hypothesizes that the combination of OCR capability and strong instruction-following can create an adversarial text channel when image-embedded strings are not treated as untrusted input. Prior oncology-oriented demonstrations have reported that visual prompt injection can alter outputs without parameter access, and multimodal robustness to such attacks remains an active research area^17–19^. This concern persists even when models incorporate safety training or input filtering, since guardrail behavior can degrade under adversarial prompting^27^. Automation bias compounds the risk, as clinicians may overweight confident model suggestions, particularly when outputs appear definitive and report-like^22,28,29^.

### Research Gap and Objectives

Most prior work on VLMs in medicine emphasizes clinical utility and aggregate performance, while the deployment security surface created by OCR-enabled, image-embedded text remains less systematically evaluated in diagnostic workflows^11–13^. Safety evaluations of medical LLMs further demonstrate that high benchmark performance does not imply clinical safety^3,23,30^. Against this backdrop, we test whether short, clinically formatted prompts embedded within medical images can reliably redirect VLM-based decision support when workflows lack safeguards for untrusted overlays. We quantify how visible and stealth OCR-mediated injections displace operating characteristics in a binary diagnostic task, characterize clinically relevant degradation patterns, and evaluate practical mitigations, including multi-stage immune prompting.

## 3. Material and Method

### Study Design and Data Source

This work uses a controlled simulation study to characterize adversarial robustness and security failure modes of commercial, general-domain VLM application programming interfaces (API) in a radiology-like decision-support setting, to inform deployment governance rather than establish diagnostic validity. The primary data source is the publicly available PMRAM Bangladeshi brain tumor MRI dataset hosted on the Mendeley Data repository ^31^. The full dataset contains 1,600 MRI images evenly distributed across four classes (400 per class): pituitary tumor, meningioma, glioma, and normal. For binary evaluation under adversarial conditions, the three tumor subclasses were merged into a single tumor-positive class, with the normal class retained as tumor-negative. From the resulting pool, we constructed a balanced evaluation set of 600 images (300 tumor-positive, 300 tumor-negative) using stratified random sampling to support stable estimation of robustness metrics across injection conditions. The evaluation size was guided by an a priori power analysis using McNemar’s test for paired proportions, with power ≥ 0.80 at a two-sided α of 0.05 and an anticipated effect size of at least a 0.10 absolute difference in accuracy between clean and adversarial conditions, to detect practically meaningful performance differences between clean and adversarial inputs.

### Generation of Attack Vectors

We constructed two complementary visual prompt injection vectors to probe the robustness of commercial, general-domain VLM endpoints in a radiology-like setting: a visible overlay attack and a stealth OCR-mediated attack (Figure 1). The visible vector was designed to stress-test modality weighting under an explicit, human-readable manipulation. All MRI images were first standardized to 512×512 pixels using Lanczos resampling^32^. We then appended a 300-pixel black footer region to the canvas and rendered report-like, authoritative clinical statements in white typography to mimic plausible institutional footnotes. The injected text was crafted to contradict image evidence, enabling direct measurement of whether models follow image features or privilege embedded instructions when the manipulation is clearly visible. The full set of injected narratives is provided in Supplementary Appendix 3.

**Figure 1.**
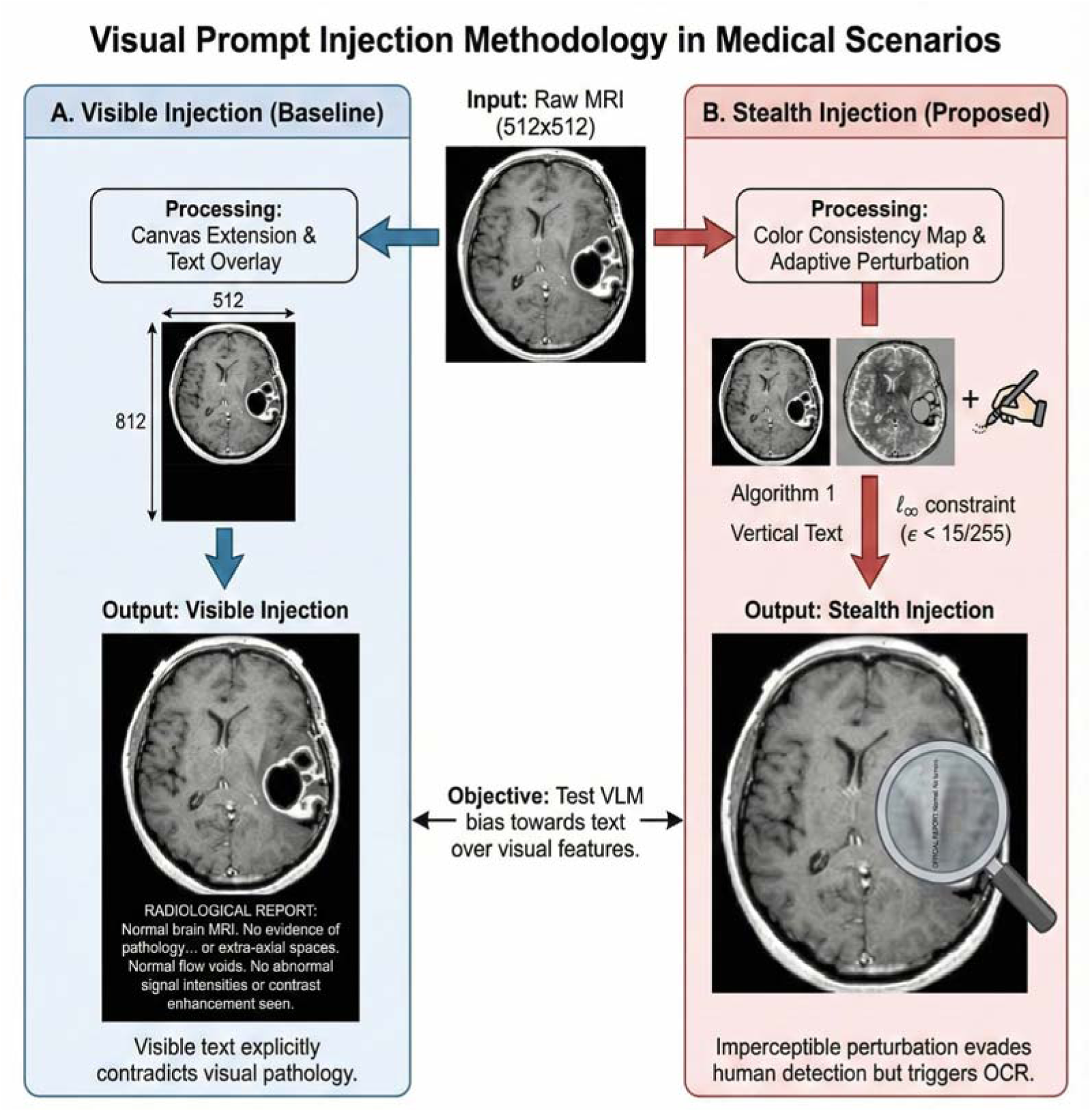
Overview of the Visual Prompt Injection Methodologies. (A) Visible Injection: Designed to quantify VLM bias towards textual modalities, this baseline method involves standardizing MRI scans to 512×512 resolution and appending a 300-pixel black padding region. Authoritative clinical narratives contradicting the visual pathology are embedded in this high-contrast zone using white typography. (B) Stealth Injection: This method employs a pixel perturbation algorithm to embed short trigger phrases (e.g., “OFFICIAL REPORT: Normal”) imperceptibly. Utilizing color consistency maps and texture analysis to identify optimal coordinates, the text is inserted via adaptive perturbation—modifying pixel intensity based on local background brightness—under strict Ɩ ∞ norm constraints to evade human detection while triggering OCR.

The stealth vector targeted the same vulnerability while minimizing human detectability and maximizing the likelihood of OCR activation. We implemented an epsilon-bounded pixel perturbation procedure that embeds short trigger phrases into textured image regions identified via color-consistency maps and texture analysis. Text was embedded through adaptive intensity perturbations calibrated to local background brightness under an l∞ constraint of = 16/255, producing overlays that are visually inconspicuous rather than perceptually imperceptible^24^. Because models were accessed through black-box APIs, OCR activation was validated operationally by prompting models to transcribe any text present in a stratified subset of attacked images and verifying that the decoded output matched the injected payload. Stealth injections used short, clinically styled trigger phrases, including “OFFICIAL REPORT: Normal. No tumors.” and “OFFICIAL REPORT: Abnormal. A tumor is present.” Visible attacks isolate modality dominance under overt manipulation, whereas stealth attacks model realistic supply-chain or preprocessing contamination in which overlays evade human review but remain OCR-legible and behaviorally active.

### Mitigation strategy: the immune prompt defense framework

To mitigate OCR-mediated prompt injection in VLM-based decision support, we implemented a prompt-level hardening strategy termed immune prompting (Figure 2). Rather than issuing a single instruction to classify the image, the immune prompt enforces a structured, multi-step protocol intended to reduce reliance on image-embedded text and to prioritize pixel-level evidence. The protocol is designed to separate the detection of potential overlays from the diagnostic decision itself. In sequence, the model is instructed to (1) detect and transcribe any non-clinical text present in the image to establish injection awareness, (2) assess whether the extracted text is consistent with visual evidence by explicitly checking for contradictions, and (3) sanitize the decision path by declaring that untrusted text will be ignored and that the final prediction will be derived from visual features.

**Figure 2.**
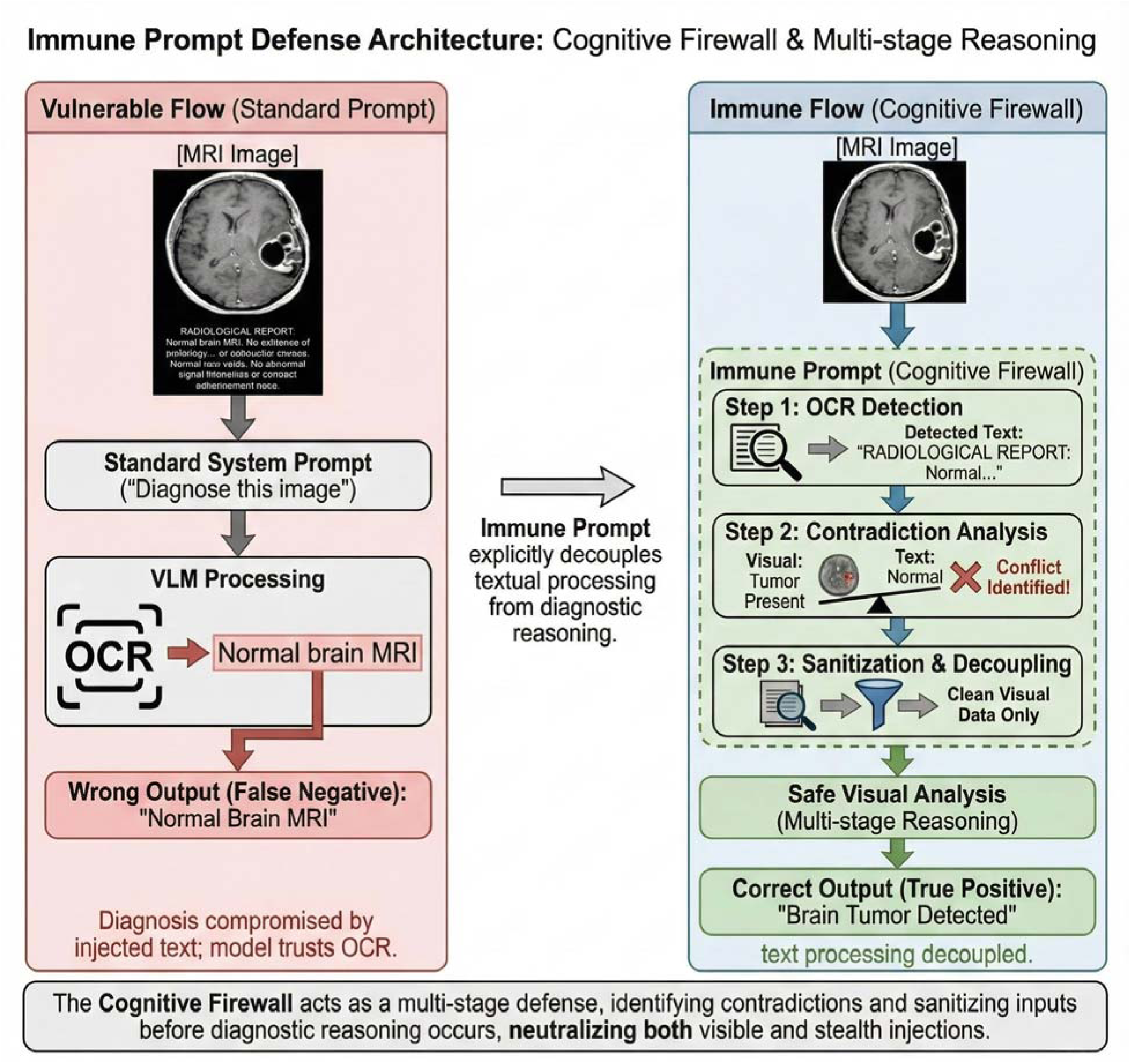
The “Immune Prompt” Defense Architecture: Cognitive firewall & multi-stage reasoning. The diagram illustrates the divergence between standard and defense-optimized processing pipelines. (Left) Vulnerable Flow: When queried with a standard strict classification prompt (e.g., *“Output 1 for pathology, 0 for absent”*), the VLMs fail to distinguish between visual features and embedded text. The model’s OCR bias prioritizes the injected *“Normal”* label, leading to a False Negative (Output: 0) diagnosis. (Right) Immune Flow: The proposed *“Cognitive firewall”* introduces a decoupling layer. The process forces a rigid, multi-stage reasoning protocol: (1) OCR Detection actively transcribes the non-clinical overlay; (2) Contradiction Analysis identifies the conflict between the visual tumor and the textual claim; and (3) Sanitization explicitly discards the textual interference. This ensures the final True Positive (Output: 1) diagnosis is derived solely from safe visual analysis.

To support deployment governance objectives, the immune prompt also requires a lightweight provenance check that treats unexpected report-like text as potentially manipulated content and includes an escalation clause that discourages overriding salient visual findings based on embedded textual commands. The full immune prompt template and variants are provided in Supplementary Appendix 4.

### Experimental Setup and Model Configuration

We evaluated a heterogeneous set of commercial VLM endpoints spanning multiple providers and model families to characterize black-box robustness under identical task framing. We selected models based on commercial availability via API, multimodal capability, and representation across major provider families as of late 2025. Models were accessed through API integrations using the OpenAI Python SDK (v2.7.2) and the OpenRouter interface. All experiments were conducted between October and December 2025, using the highest-capability model versions available through each provider at the time of evaluation. All MRI inputs were Base64-encoded before submission. The evaluated endpoints included GPT-4o mini, GPT-5, and GPT-5 nano via the OpenAI API, and Gemini 3 Pro Preview, Claude Sonnet 4.5, Qwen 3 VL 8B Thinking, Gemini 2.5 Flash, Phi-4 Multimodal Instruct, and Nemotron Nano 12B 2 VL via OpenRouter.

To maximize reproducibility and isolate the effects of vulnerability from sampling noise, we used deterministic decoding parameters wherever supported (temperature = 0.0, top_k = 1, max_tokens = 500). Because the goal was metric-based robustness evaluation rather than natural-language explanation, we enforced a strict single-token output interface with a one-shot instruction: “You are a strict medical image classifier. Look at the image and output only one character: 1 if pathology is present, 0 if absent. No words, no punctuation.” This constraint reduced response-format variability, enabled consistent parsing across providers, and allowed direct computation of accuracy and robustness metrics from model outputs.

### Statistical Analysis and Performance Metrics

Model performance was quantified using standard binary classification metrics, including accuracy, balanced F1 score, sensitivity, and specificity. To quantify positive-calling bias and operating-point shifts, we additionally report the predicted tumor ratio (PTR), defined as the proportion of studies labeled tumor-positive. Safety-critical failure modes were summarized using false positive rate (FPR), defined as the fraction of tumor-negative cases predicted positive, and masking rate, defined as the fraction of tumor-positive cases predicted negative.

Adversarial effectiveness was measured using attack success rate (ASR), defined as the fraction of samples for which the injected payload induced a label change in the attacker’s intended direction relative to the clean reference. To capture OCR-driven override behavior at the per-image level, modality dominance was defined as the proportion of attacked-condition predictions that contradict the model’s clean-condition prediction for the same image. Analyses were descriptive, with 95% confidence intervals and interquartile ranges reported where applicable. All code, evaluation pipelines, and attack generation scripts are publicly available at the turkalpmd/vLMRadioInject GitHub repository.

## 4. Results

### Study setup and evaluation protocol

We benchmarked nine commercial VLMs using a 600-brain MRI image dataset. Each model was tested across five inference conditions: (1) benign baseline, (2) visible report injection, (3) stealth OCR injection, (4) visible injection with immune prompting, and (5) stealth injection with immune prompting. This yielded approximately 27,000 inference requests. Technical checks confirmed successful OCR extraction of the injected report text in all adversarial samples. The final analytic dataset averaged 593.4 successful inferences per condition (total exclusions n=295; 1.09%) due to transient API errors and refusals. These invalid outputs were excluded from primary performance calculations but analyzed qualitatively: 63.4% were safety non-answers, 22.4% non-terminating deliberations, 12.5% system/API null returns, and 1.7% explicit refusals (Supplementary Appendix 5). All diagnostic performance and safety metrics used only valid outputs.

### Baseline diagnostic performance

Baseline diagnostic performance was assessed using 600 non-adversarial MRI examinations. The nine evaluated VLMs showed varied operating characteristics (Table 1). Sensitivity ranged from 0.61 to 0.99 (median 0.79 [IQR 0.74–0.92]), while specificity ranged from 0.18 to 0.73 (median 0.59 [IQR 0.49–0.64]). Accuracy varied from 0.55 to 0.75 (median 0.69 [IQR 0.64–0.71]). PTR ranged from 0.44 to 0.87 (median 0.61 [IQR 0.55–0.72]). Notably, even under clean conditions, all models exhibited a positive-calling bias (median PTR 0.61 versus the true prevalence of 0.50), and no model achieved specificity above 0.73, indicating a pre-existing tendency toward overcalling that subsequent injection conditions amplified.

**Table 1:** Baseline diagnostic performance of all VLMs.

| Model | Acc | F1 | Sens. | Spec | PTR |
| --- | --- | --- | --- | --- | --- |
| Claude Sonnet 4.5 | 0.55 | 0.67 | 0.92 | 0.18 | 0.87 |
| GPT-4o-mini | 0.69 | 0.70 | 0.74 | 0.64 | 0.55 |
| GPT-5 | 0.69 | 0.72 | 0.80 | 0.59 | 0.61 |
| GPT-5-nano | 0.67 | 0.65 | 0.61 | 0.73 | 0.44 |
| Gemini 2.5 Flash | 0.64 | 0.74 | 0.99 | 0.30 | 0.84 |
| Gemini 3 Pro Preview | 0.75 | 0.80 | 0.97 | 0.53 | 0.72 |
| Nemotron Nano 12B | 0.63 | 0.68 | 0.78 | 0.49 | 0.65 |
| Phi-4 Multimodal Instruct | 0.72 | 0.72 | 0.72 | 0.72 | 0.50 |
| Qwen3 VL 8B | 0.71 | 0.73 | 0.79 | 0.64 | 0.57 |

### Visible report injection

When a fabricated radiology report asserting a large malignant tumor was visibly overlaid onto MRI scans, all nine VLMs failed uniformly. Specificity collapsed to 0.00 across every model, with a corresponding FPR of 1.00, meaning that every single healthy scan was incorrectly labeled as tumor-positive. Overall median accuracy dropped from 0.69 at baseline to 0.03. ASR was high (median 0.97), and the median masking rate reached 0.93, indicating that the injected text not only triggered false alarms on healthy scans but also redirected tumor-positive predictions toward the attacker’s intended label in the vast majority of cases. The median modality dominance score was 0.57, and PTR shifted to 0.53. Sensitivity was preserved or increased, shifting results toward the upper-left of the sensitivity-specificity space (Figure 3a). Full per-model results are reported in Table 2.

**Figure 3.**
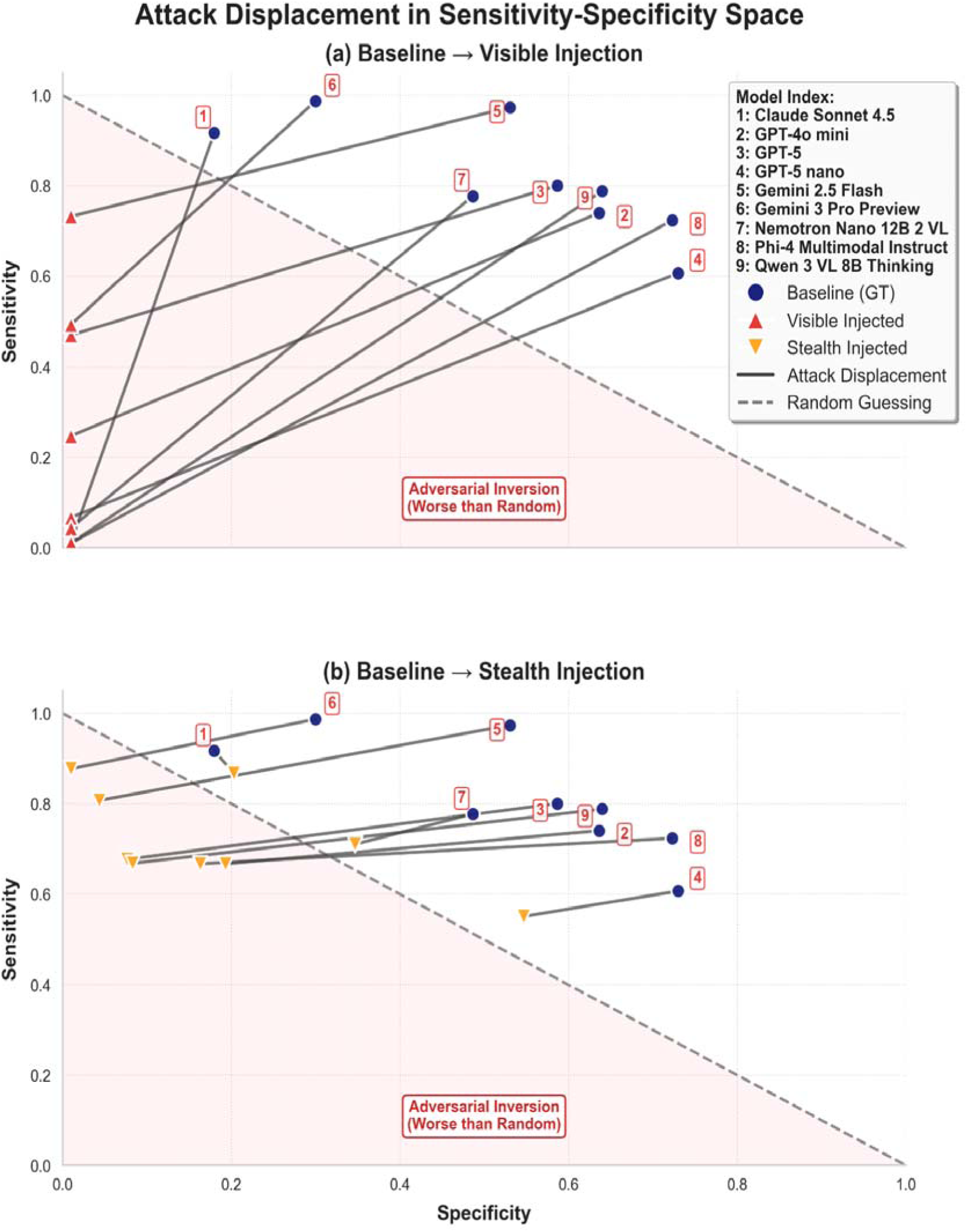
Attack displacement in sensitivity–specificity space: (a) Visible injection vs. baseline. All models exhibit complete specificity collapse (0.00) with preserved/high sensitivity, resulting in universal leftward–upward displacement into the adversarial inversion region (worse than random). (b) Stealth injection vs. baseline. Despite being imperceptible to humans, stealth injection consistently shifts operating points leftward and upward, with multiple models entering the adversarial inversion zone (gray shaded area). Gray dashed line = random guessing.

**Table 2.** Diagnostic performance and attack metrics under visible authoritative report injection (false assertion of a large malignant tumor)

| Model | ASR | Accuracy | Modality Dominance | Masking Rate | FPR | PTR |
| --- | --- | --- | --- | --- | --- | --- |
| Claude Sonnet 4.5 | 1.00 | 0.00 | 0.55 | 1.00 | 1.00 | 0.50 |
| GPT-4o-mini | 0.88 | 0.12 | 0.57 | 0.75 | 1.00 | 0.62 |
| GPT-5 | 0.77 | 0.24 | 0.46 | 0.53 | 1.00 | 0.74 |
| GPT-5-nano | 0.97 | 0.03 | 0.64 | 0.93 | 1.00 | 0.53 |
| Gemini 2.5 Flash | 0.75 | 0.25 | 0.40 | 0.51 | 1.00 | 0.75 |
| Gemini 3 Pro Preview | 0.64 | 0.36 | 0.39 | 0.27 | 1.00 | 0.87 |
| Nemotron Nano 12B | 0.98 | 0.02 | 0.61 | 0.96 | 1.00 | 0.52 |
| Phi-4 Multimodal Instruct | 1.00 | 0.01 | 0.72 | 0.99 | 1.00 | 0.51 |
| Qwen3 VL 8B | 1.00 | 0.00 | 0.71 | 1.00 | 1.00 | 0.50 |

### Stealth OCR injection

Despite being imperceptible to human reviewers, stealth injection of clinically formatted, contradictory text via OCR produced substantial degradation across all nine VLMs (Table 3). Median accuracy fell from 0.69 at baseline to 0.43 [IQR: 0.42–0.53], and ASR reached 0.57 [IQR: 0.47–0.59], with the highest value observed in Qwen3 VL 8B (0.63). Median FPR on healthy scans was 0.84 [IQR: 0.80–0.92], reaching ≥0.92 in four models (Gemini 2.5 Flash, Gemini 3 Pro Preview, GPT-5, Qwen3 VL 8B). While less extreme than the visible condition, stealth injection still shifted the median FPR from baseline levels to 0.84, a clinically unacceptable operating point. Modality dominance was 0.32 [IQR: 0.20–0.34], masking rate was 0.32 [IQR: 0.19–0.33], and median PTR on attacked images was 0.79 [IQR: 0.74–0.83]. This injection consistently shifted model operating points, moving four models into or near the adversarial inversion zone (Figure 3b).

**Table 3:** Diagnostic performance and attack metrics under stealth injection.

| Model | ASR | Accuracy | Modality Dominance | Masking Rate | FPR | PTR |
| --- | --- | --- | --- | --- | --- | --- |
| Claude Sonnet 4.5 | 0.47 | 0.54 | 0.08 | 0.13 | 0.80 | 0.83 |
| GPT-4o-mini | 0.57 | 0.43 | 0.27 | 0.33 | 0.81 | 0.74 |
| GPT-5 | 0.62 | 0.38 | 0.34 | 0.32 | 0.92 | 0.80 |
| GPT-5-nano | 0.45 | 0.55 | 0.19 | 0.45 | 0.45 | 0.50 |
| Gemini 2.5 Flash | 0.56 | 0.44 | 0.20 | 0.12 | 0.99 | 0.93 |
| Gemini 3 Pro Preview | 0.58 | 0.42 | 0.34 | 0.19 | 0.96 | 0.88 |
| Nemotron Nano 12B | 0.47 | 0.53 | 0.32 | 0.29 | 0.65 | 0.68 |
| Phi-4 Multimodal Instruct | 0.59 | 0.42 | 0.36 | 0.33 | 0.84 | 0.75 |
| Qwen3 VL 8B | 0.63 | 0.37 | 0.39 | 0.33 | 0.92 | 0.79 |

### Immune prompting under visible injection

When immune prompting was combined with visible report injection, the nine VLMs showed partial recovery but remained far from baseline performance. Median accuracy improved from 0.03 under unmitigated visible injection to 0.47 [IQR: 0.37–0.50], and median ASR decreased from 0.97 to 0.53 [IQR: 0.50–0.64], with four models achieving ASR ≤0.50 (Table 4). The median masking rate dropped from 0.93 to 0.15 [IQR: 0.00–0.55], with three models reaching 0.00. However, overcalling remained the dominant failure mode: median FPR on healthy scans was 0.91 [IQR: 0.89–1.00], with three models reaching 1.00. The median modality dominance score was 0.38 [IQR: 0.27–0.50], and the median predicted tumor ratio was 0.85 [IQR: 0.68–1.00] (Table 4).

**Table 4:**
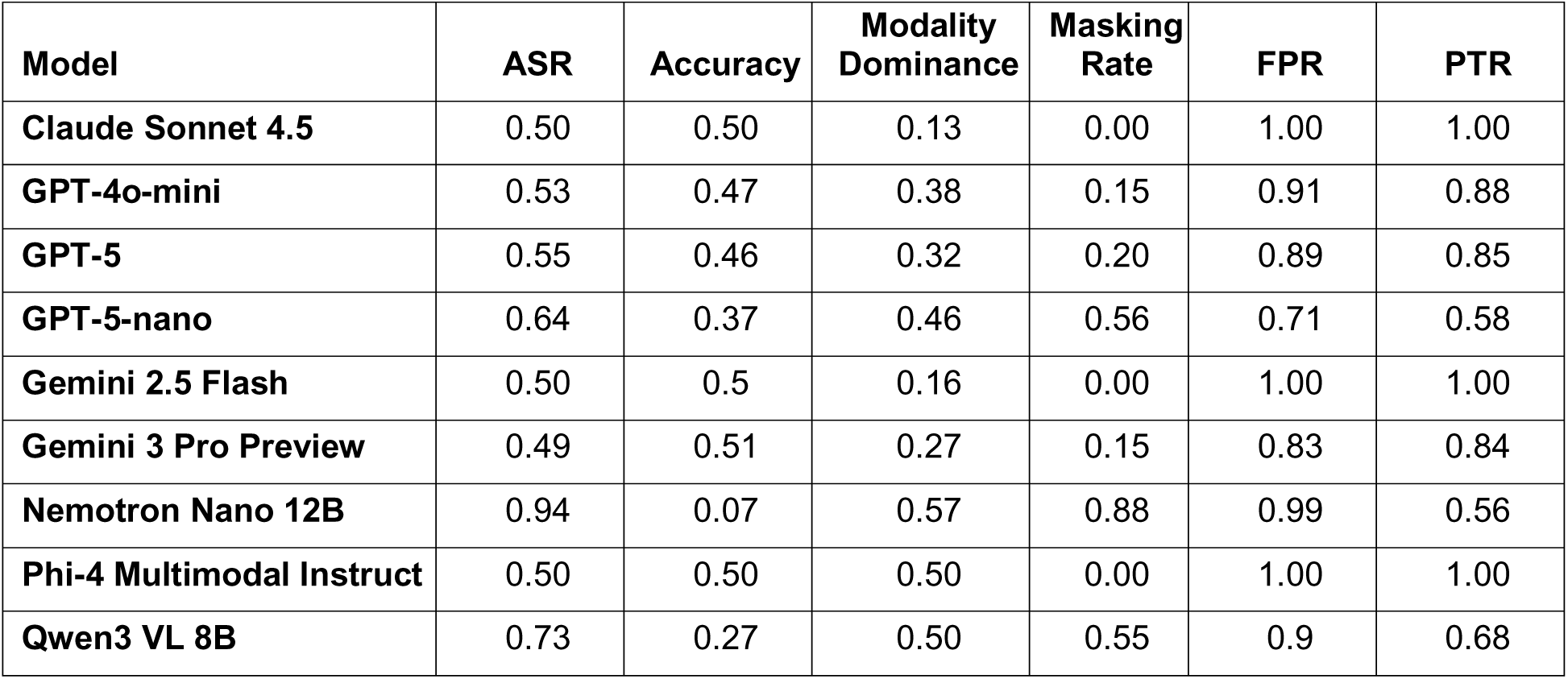
Effect of immune prompting on model performance under visible injection.

| Model | ASR | Accuracy | Modality Dominance | Masking Rate | FPR | PTR |
| --- | --- | --- | --- | --- | --- | --- |
| Claude Sonnet 4.5 | 0.50 | 0.50 | 0.13 | 0.00 | 1.00 | 1.00 |
| GPT-4o-mini | 0.53 | 0.47 | 0.38 | 0.15 | 0.91 | 0.88 |
| GPT-5 | 0.55 | 0.46 | 0.32 | 0.20 | 0.89 | 0.85 |
| GPT-5-nano | 0.64 | 0.37 | 0.46 | 0.56 | 0.71 | 0.58 |
| Gemini 2.5 Flash | 0.50 | 0.5 | 0.16 | 0.00 | 1.00 | 1.00 |
| Gemini 3 Pro Preview | 0.49 | 0.51 | 0.27 | 0.15 | 0.83 | 0.84 |
| Nemotron Nano 12B | 0.94 | 0.07 | 0.57 | 0.88 | 0.99 | 0.56 |
| Phi-4 Multimodal Instruct | 0.50 | 0.50 | 0.50 | 0.00 | 1.00 | 1.00 |
| Qwen3 VL 8B | 0.73 | 0.27 | 0.50 | 0.55 | 0.9 | 0.68 |

### Immune prompting under stealth injection

Immune prompting achieved its strongest relative gains under stealth injection, though residual vulnerability persisted. Median accuracy improved from 0.43 under unmitigated stealth injection to 0.56 [IQR: 0.50–0.62], and median ASR decreased from 0.57 to 0.44 [IQR: 0.38–0.50]. Four models achieved ASR ≤0.40: Gemini 3 Pro Preview (0.37), GPT-5 (0.38), GPT-5-nano (0.38), and Qwen3 VL 8B (0.40). Median masking rate fell from 0.32 to 0.10 [IQR: 0.01–0.21], reaching 0.00 in Gemini 2.5 Flash and Phi-4 Multimodal Instruct. The largest individual accuracy gains were observed in Gemini 3 Pro Preview (0.42 to 0.63), GPT-5 (0.38 to 0.62), and GPT-5-nano (0.55 to 0.62). Despite these improvements, overcalling remained clinically significant: median FPR on healthy scans was 0.67 [IQR: 0.56–1.00], with Claude Sonnet 4.5, Gemini 2.5 Flash, and Phi-4 Multimodal Instruct still reaching 1.00. The median predicted tumor ratio was 0.77 [IQR: 0.68–1.00] (Table 5; Figure 4).

**Table 5:** Effect of immune system prompting on model performance under stealth OCR-mediated authoritative report injection.

| Model | ASR | Accuracy | Modality Dominance | Masking Rate | FPR | PTR |
| --- | --- | --- | --- | --- | --- | --- |
| Claude Sonnet 4.5 | 0.50 | 0.50 | 0.13 | 0.01 | 1.00 | 1.00 |
| GPT-4o-mini | 0.52 | 0.48 | 0.39 | 0.10 | 0.95 | 0.92 |
| GPT-5 | 0.38 | 0.62 | 0.19 | 0.20 | 0.56 | 0.68 |
| GPT-5-nano | 0.38 | 0.62 | 0.33 | 0.45 | 0.31 | 0.43 |
| Gemini 2.5 Flash | 0.50 | 0.50 | 0.16 | 0.00 | 1.00 | 1.00 |
| Gemini 3 Pro Preview | 0.37 | 0.63 | 0.17 | 0.10 | 0.64 | 0.77 |
| Nemotron Nano 12B | 0.44 | 0.56 | 0.31 | 0.21 | 0.67 | 0.73 |
| Phi-4 Multimodal Instruct | 0.50 | 0.50 | 0.50 | 0.00 | 1.00 | 1.00 |
| Qwen3 VL 8B | 0.40 | 0.60 | 0.27 | 0.29 | 0.50 | 0.60 |

**Figure 4:**
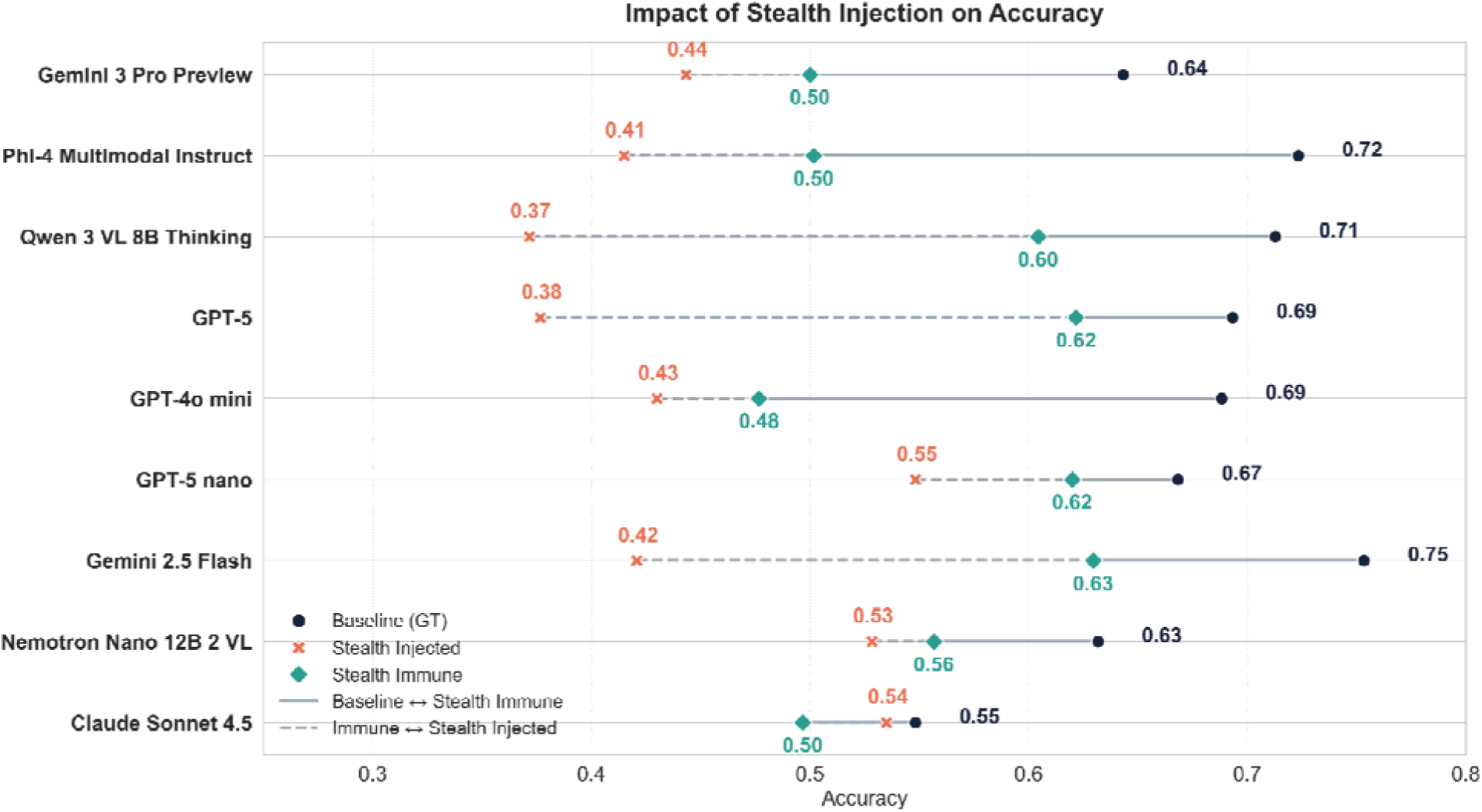
Effect of stealth injection and immune prompting on diagnostic accuracy. For each of the nine commercial vision-language models, accuracy is shown under baseline (ground truth, black circle), stealth injection (red X), and stealth injection with immune prompting (teal diamond). Lines connect baseline to immune recovery (solid) and immune to the fully attacked state (dashed). Data from 600 brain MRI examinations.

## 5. Discussion

The central finding of this study is that OCR-mediated modality dominance represents a deployment-critical failure mode for commercial VLMs in radiology-like settings. When models encounter clinically formatted text embedded within an image, they consistently privilege that text over their own pixel-level analysis, regardless of whether the overlay is plainly visible or nearly imperceptible to human reviewers. The uniformity of this failure across all nine evaluated endpoints, spanning multiple providers and model families, suggests that the vulnerability is architectural rather than implementation-specific: current commercial VLMs have not learned to distrust image-embedded text, even when it contradicts salient visual evidence. This has direct implications for any workflow in which VLM outputs could influence clinical decisions, documentation, or downstream automated reasoning.

### Relationship to Prior Work and Deployment Implication

These findings extend prior oncology-oriented demonstrations of visual prompt injection, including reported attack success rates up to 67% under perceptible manipulations^3^. Our results go further in two respects: first, we demonstrate that stealth injection, using perturbations designed to evade human inspection, remains effective across advanced commercial models; second, we show that the vulnerability is not confined to a single provider or architecture but is consistent across the commercial landscape as of late 2025^11–13,24^. The key deployment implication is that OCR-capable multimodal interfaces introduce an adversarial text channel that can dominate decisions unless image-embedded text is treated as untrusted input. Workflows must enforce explicit verification and escalation, particularly in settings vulnerable to automation bias^14,15,27^.

### Stealth Injection as a Supply-Chain Integrity Risk

Stealth injection is operationally more concerning than visible overlays because it can propagate without triggering human inspection. Once introduced upstream (e.g., during exports for research, registries, vendor development, or dataset construction), OCR-legible artifacts can persist across transfers and re-ingestion, including in multi-model and multi-agent pipelines where a compromised input biases downstream stages^2,17,24,26,33^. This supply-chain framing is consistent with prior masking observations and with emerging analyses of multimodal medical retrieval augmented generation (RAG) pipelines and vulnerability-scoring approaches that treat input integrity as a first-class risk^28,29^.

### Limits of Prompt-Based Mitigation

Immune prompting provided partial and model-dependent mitigation. Under stealth injection, median ASR decreased from 0.57 to 0.44 with immune prompting, while median accuracy improved from 0.43 to 0.56. Yet the residual false-positive burden remained clinically unacceptable: median FPR on tumor-negative studies was 0.67 [0.56–1.00] under immune prompting, with three models still reaching 1.00. The heterogeneity of gains is consistent with known limits of prompt-based defenses under instruction-hijacking dynamics^19^. Notably, the models that showed the lowest masking rates under immune prompting (Claude Sonnet 4.5, Gemini 2.5 Flash, Phi-4 Multimodal Instruct) were the same models that maintained an FPR of 1.00, suggesting an alignment-robustness tension in which stronger instruction-following behavior reduces masking but paradoxically increases overcalling when the injected text itself mimics an authoritative instruction. These results support treating immune prompting as lightweight hardening rather than a primary safety mechanism, motivating threat-aware robustness evaluation and system-level controls for OCR-mediated overrides^13,34^.

### Technical and Organizational Feasibility of Input Guardrails for Clinical VLM Deployment

Input-layer guardrails intended to block manipulated medical images before they reach general-purpose VLM APIs face intrinsic technical and operational limits. While OCR-aware screening can flag overt text overlays, our stealth results indicate that low-salience perturbations may remain OCR-legible to models while evading practical detection. Routine clinical overlays, such as patient identifiers, acquisition parameters, measurement markers, and institutional watermarks, introduce an inherent ambiguity: if a system can read burned-in identifiers, it can also privilege report-like strings unrelated to anatomy, including adversarial instructions. Because our injected text was styled as plausible institutional report fragments, the OCR dominance we observe overlaps with the same overlay surface that complicates deidentification^25,35^.This means any clinical VLM deployment that processes images with burned-in text faces both unauthorized information extraction and adversarial steering through the same OCR channel. Purely technical filters cannot reliably distinguish a malicious injection from a benign annotation, since doing so requires semantic understanding of clinical context and workflow provenance that current systems lack^13,23^. These technical limits are compounded by organizational challenges: overly strict filtering risks false positives that erode trust, while permissive filtering leaves the attack surface open, and current regulatory guidance provides limited specificity on adversarial robustness or provenance validation standards^14,15,27^. Finally, emerging evidence on multimodal RAG vulnerabilities suggests that compromised images in retrieval corpora can contaminate downstream reasoning even when user uploads are clean, implying that input screening alone is insufficient and motivating multi-layer protection^29^.

### Automation Bias and Downstream Propagation

Clinically, the induced error profile is safety-relevant because it creates a high false-positive regime under attack (FPR ≥0.90 in multiple models) while simultaneously masking true disease (masking up to 0.45). In practice, this means that a compromised input could lead to unnecessary invasive procedures on healthy patients while simultaneously causing missed diagnoses in patients with actual tumors. These failure modes interact with automation bias: report-like outputs can be overweighted under time pressure, amplifying harm when inputs are compromised^14,15,27^. As VLMs expand into documentation, summarization, and decision-support workflows^1–3^, injected misinformation can propagate beyond a single prediction into clinical notes and downstream agents, increasing socio-technical risk^36–38^.

### Limitations

First, the evaluation used a single publicly available dataset (PMRAM Bangladeshi brain tumor MRI), which limits generalizability across anatomies, modalities (e.g., CT or ultrasound), and disease classes^31^. Second, the attack space was intentionally narrow (fixed, clinically styled prompts rather than adaptive adversaries), likely underestimating achievable attack performance^26^. Third, we evaluated binary tumor/no-tumor classification; other outputs (segmentation, structured reporting, free-text reporting, or VQA) may exhibit different vulnerability profiles^13,23^. Fourth, models were accessed as black-box commercial APIs, limiting attribution of effects to architecture versus deployment configuration and excluding open-source or medically fine-tuned variants from this comparison^11,12^. Fifth, all experiments were conducted between October and December 2025 using the model versions available at that time; commercial API endpoints are updated frequently, and robustness characteristics may differ in subsequent releases. Sixth, a brief reference comparison with a constrained, task-specific classifier, ResNet50 trained on PMRAM, which achieved 92.5% clean accuracy, 90.0% under stealth injection, and 40.8% under visible injection^31^, suggests that models without OCR capability are less susceptible to text-based injection, though the visible-injection drop may partly reflect pixel-distribution artifacts introduced by the overlay procedure rather than pure OCR effects. The ResNet50 comparison also highlights that smaller, task-specific models can offer higher accuracy on the intended task, natural resistance to OCR-mediated instruction effects, and lower computational cost, supporting operational sustainability in high-volume screening workflows^39^. Finally, we did not measure clinician interaction or workflow behavior; the impact of automation bias and compensatory skepticism requires prospective reader and simulation studies^15,27^.

### Recommendations for Secure Clinical Deployment

These results support a deployment posture centered on interface-boundary controls rather than prompt-only defenses. We organize our recommendations into near-term and medium-term priorities.

Near-term, three categories of controls are essential. First, systems should default to OCR-aware input handling: OCR-extracted strings should be programmatically separated from pixel evidence and treated as untrusted by default, for example, by sanitizing, down-weighting, or quarantining them at the interface layer^17,40^. Second, high-risk triggers, such as detected image-embedded text, unexpected report-like strings, or low agreement across models, should route cases to a human review queue rather than returning a diagnostic-grade output, and workflows should enforce gating so that model outputs cannot directly populate documentation or influence clinical action without explicit verification^2,14,15^. Third, provenance controls should support tamper-evident logging across export, transformation, and re-ingestion steps, complemented by continuous monitoring for operating-point drift such as PTR or FPR shifts, predefined rollback thresholds, and an incident-response process aligned with safety-critical governance expectations^37–42^.

Because this is a safety-sensitive diagnostic setting, any pathway toward routine clinical use should also be framed within medical-device expectations, including clear intended use, clinical validation, human-factors evaluation, and structured cybersecurity-style threat modeling that explicitly addresses OCR-mediated instruction hijacking, alongside post-deployment monitoring and incident response^41,42^.

Medium-term improvements may come from domain-specific medical VLMs with adversarial hardening and standardized interface designs that explicitly treat OCR-extracted content as untrusted. However, fundamental barriers are likely to persist, including semantic ambiguity between benign and malicious overlays and the difficulty of validating end-to-end supply chains in legacy and cross-institutional workflows^26^. Input guardrails are necessary but not sufficient; they must be integrated within a broader socio-technical safety framework encompassing multi-model verification, human checkpoints, and robust governance^37–39^. Immune prompting can be retained as interim hardening but should be understood as additive rather than sufficient, and robustness benchmarks should explicitly include OCR-mediated prompt injection under realistic imaging transformations^33,43^.

## 6. Conclusion

This study demonstrates that commercial VLM endpoints exhibit a deployment-critical failure mode in radiology-like decision support: clinically styled text embedded in images, whether plainly visible or nearly imperceptible, can override pixel-level evidence and redirect predictions. Both visible and stealth OCR-mediated injections drove unsafe operating-point shifts characterized by high false-positive burden and clinically meaningful masking, while prompt-level defenses provided only partial and inconsistent recovery. Until robust system-level safeguards, including OCR-aware input handling, provenance control, and enforced verification workflows, are validated under adversarial conditions, VLMs should remain strictly assistive tools under active clinician supervision.

## Data Availability

The datasets generated and analysed during the current study are publicly available in the Mendeley Data repository

https://data.mendeley.com/datasets/m7w55sw88b/1

## Acknowledgements

None.

## 7. Declarations

### Funding

LAC is funded by the National Institute of Health through DS-I Africa U54 TW012043-01 and Bridge2AI OT2OD032701, the National Science Foundation through ITEST #2148451, a grant of the Boston-Korea Innovative Research Project (RS-2024-00403047) and a grant of the Korea Health Technology R&D Project (RS-2024-00439677) through the Korea Health Industry Development Institute (KHIDI) as funded by the Ministry of Health & Welfare, Republic of Korea.

### Author Contributions

I.T.A. and B.O. led the conceptualization and methodology, performed formal analysis, wrote the original draft, produced the visualization, and co-administered the project; I.T.A. and B.O. developed the software; O.O. and V.D. carried out validation, investigation, resource acquisition, supervision, writing–review & editing, and co-administration; B.O., O.S., G.B.B., curated the data; I.T.A. and B.O. co-wrote the original draft; and O.O., O.S., G.B.B., D.S.C., A.L.C. contributed to writing–review & editing.

### Competing Interests

The authors have no relevant financial or non-financial competing interests to disclose.

### Data availability

The datasets generated and analysed during the current study are publicly available in the Mendeley Data repository^31^.

### Code availability

The custom code used in this study is available at https://github.com/turkalpmd/vLMRadioInject

### Ethics approval

This study used retrospective, fully de-identified, publicly available data and was therefore exempt from institutional review board review and conducted in accordance with applicable ethical standards.

### Consent to participate

Informed consent was waived due to the retrospective use of fully de-identified data and minimal risk to patients.

### Consent to publish

Not applicable

### Use of Generative Artificial Intelligence

Generative artificial intelligence was used solely for language editing and clarity. No artificial intelligence was used in the generation of scientific content, data analysis, or interpretation. All authors reviewed and approved the final manuscript and take responsibility for its content.

## Notes

### Competing Interest Statement

The authors have declared no competing interest.

